# Tumor-Colonizing Microbiota Distinguish Early- and Late-Onset Colorectal Cancer in a Hispanic/Latino Patient Cohort

**DOI:** 10.64898/2026.07.19.26358429

**Authors:** Sophia Manjarrez, Fernando C Diaz, Francisco G. Carranza, Brigette Waldrup, Maria Ninova, Enrique Velazquez-Villarreal

## Abstract

**Background:** Early-onset colorectal cancer (EOCRC) is increasing globally, particularly among Hispanic/Latino (H/L) populations, yet the contribution of tumor-colonizing microbiota to age-associated colorectal cancer (CRC) biology remains poorly understood. Most microbiome studies have focused on fecal communities or non-Hispanic populations, leaving the intratumoral microbial landscape of H/L patients largely unexplored.

**Methods:** We performed an exploratory characterization of tumor-colonizing microbiota using whole-exome sequencing (WES) data from four primary colorectal tumors obtained from H/L patients treated at City of Hope, including two EOCRC (<50 years) and two late-onset colorectal cancer (LOCRC; ≥50 years) cases. Following removal of host-derived sequences, microbial taxonomic profiling was conducted at the family, genus, and species levels, and microbial metabolic pathways were inferred. Clinical and pathological data were integrated to evaluate age-associated differences in microbial composition and predicted function.

**Results:** Family-, genus-, and species-level analyses consistently demonstrated greater microbial diversity in LOCRC than EOCRC. LOCRC contained more than twice the number of unique bacterial families, nearly three times as many unique genera, and more than twice as many unique bacterial species. A conserved core microbiota, including Fusobacteriaceae, Prevotellaceae, Fusobacterium, and Prevotella, was identified across both age groups, whereas LOCRC was enriched in CRC-associated taxa including Fusobacterium nucleatum, Bacteroides fragilis, Parvimonas micra, Porphyromonas asaccharolytica, and Dialister pneumosintes. Species-level analyses revealed only a single shared bacterial species between EOCRC and LOCRC, indicating progressive microbial divergence with increasing taxonomic resolution. In contrast, functional profiling identified 11 predicted microbial metabolic pathways, of which nine were shared between age groups, two were unique to EOCRC, and none were exclusive to LOCRC. Core metabolic pathways involved in energy metabolism, amino acid biosynthesis, phospholipid metabolism, and central carbon metabolism exhibited comparable abundance across both groups, demonstrating substantial functional conservation despite pronounced taxonomic differences.

**Conclusions:** Tumor-colonizing microbiota differ markedly between EOCRC and LOCRC in H/L patients, with late-onset tumors exhibiting substantially greater microbial richness and taxonomic complexity. Despite these compositional differences, microbial metabolic functions remain largely conserved, supporting the concept of functional redundancy within the colorectal tumor microenvironment (TME). Although exploratory, this proof-of-concept study provides one of the first characterizations of intratumoral microbiota in H/L EOCRC and establishes a foundation for larger multi-omics investigations aimed at identifying microbiome-based biomarkers and therapeutic targets for precision oncology.

## 1. Introduction

Colorectal cancer (CRC) is the third most frequently diagnosed malignancy and the second leading cause of cancer-related mortality worldwide, accounting for more than 1.9 million new cases annually. Although the overall incidence of CRC has declined in countries with established screening programs, the incidence of early-onset colorectal cancer (EOCRC), defined as CRC diagnosed before 50 years of age, continues to rise globally, particularly among Hispanic/Latino (H/L) populations, highlighting an urgent need to elucidate the biological mechanisms underlying this emerging public health challenge. [1–5]

The gut microbiome has emerged as a critical component of colorectal carcinogenesis and a dynamic regulator of the tumor microenvironment (TME). Accumulating evidence demonstrates that tumor-associated microbial dysbiosis contributes to malignant transformation through multiple mechanisms, including chronic inflammation, epithelial barrier disruption, immune modulation, genotoxicity, and microbial metabolite production. [1–4] Representative tumor-promoting microorganisms, including Fusobacterium nucleatum, enterotoxigenic Bacteroides fragilis (ETBF), and colibactin-producing Escherichia coli, have been implicated in DNA damage, immune evasion, and activation of oncogenic signaling pathways, whereas depletion of short-chain fatty acid-producing commensals further promotes tumor progression. [2–10]

Recent advances in shotgun metagenomics, metabolomics, and multi-omics integration have established reproducible microbial signatures associated with CRC development, progression, prognosis, and therapeutic response. [11–13,14–21] However, most studies have focused on fecal microbiota or predominantly European and Asian populations, limiting the understanding of tumor-colonizing microbial communities across diverse ancestral groups. Furthermore, although several investigations have demonstrated microbiome differences between EOCRC and LOCRC, these studies remain limited by relatively small sample sizes, heterogeneous methodologies, lack of clinicopathologic and molecular integration, and underrepresentation of H/L patients. [22–25] Consequently, whether age-specific tumor-resident microbial ecosystems contribute to the unique biology of EOCRC in H/L populations remains largely unknown.

H/L individuals experience a disproportionate burden of EOCRC and exhibit distinct genetic ancestry [26–29], environmental exposures, dietary patterns, and socioeconomic determinants of health that may shape host-microbiome interactions. These factors likely influence microbial composition and function within the TME, potentially contributing to differences in tumor biology, immune landscapes, therapeutic response, and clinical outcomes. Despite growing recognition of these disparities, comprehensive characterization of tumor-colonizing microbiota in H/L EOCRC has not been systematically investigated.

Here, we performed an integrative characterization of tumor-colonizing microbiota in H/L patients with EOCRC and LOCRC, leveraging high-throughput sequencing and comprehensive clinical and molecular annotation. We sought to identify age-associated microbial signatures, evaluate their relationships with clinicopathologic characteristics and genomic alterations, and define microbial communities that distinguish EOCRC from LOCRC. Our findings provide new insights into the contribution of the tumor microbiome to age-specific CRC biology and establish a foundation for microbiome-informed precision oncology in H/L populations.

## 2. Results

### 2.1. Demographic Characteristics of the H/L CRC Cohort

The study cohort comprised 4 H/L patients with primary CRC, including 2 patients with EOCRC (<50 years) and 2 with LOCRC (≥50 years). The balanced representation of EOCRC and LOCRC, together with the inclusion of both sexes and multiple disease stages, provided a unique opportunity to investigate age-associated differences in tumor-colonizing microbiota within an exclusively H/L population. This design minimizes potential confounding by ancestry and establishes a foundation for identifying microbial signatures associated with EOCRC versus LOCRC.

### 2.2. Identification of Structural Tumor-Colonizing Microbiota in EOCRC and LOCRC

#### 2.2.1. Family-Level Taxonomic Composition

To characterize the structural composition of tumor-colonizing microbiota in H/L CRC, we analyzed whole-exome sequencing (WES) data from primary colorectal tumors, including EOCRC and LOCRC cases. Following removal of host-derived reads, microbial taxonomic profiling was performed, enabling identification of bacterial taxa directly from tumor-derived sequencing data (Fig. 1A,B). Although WES is primarily designed for host genomic analyses, sufficient microbial signal was detected to permit reliable taxonomic classification at the family level.

**Figure 1.**
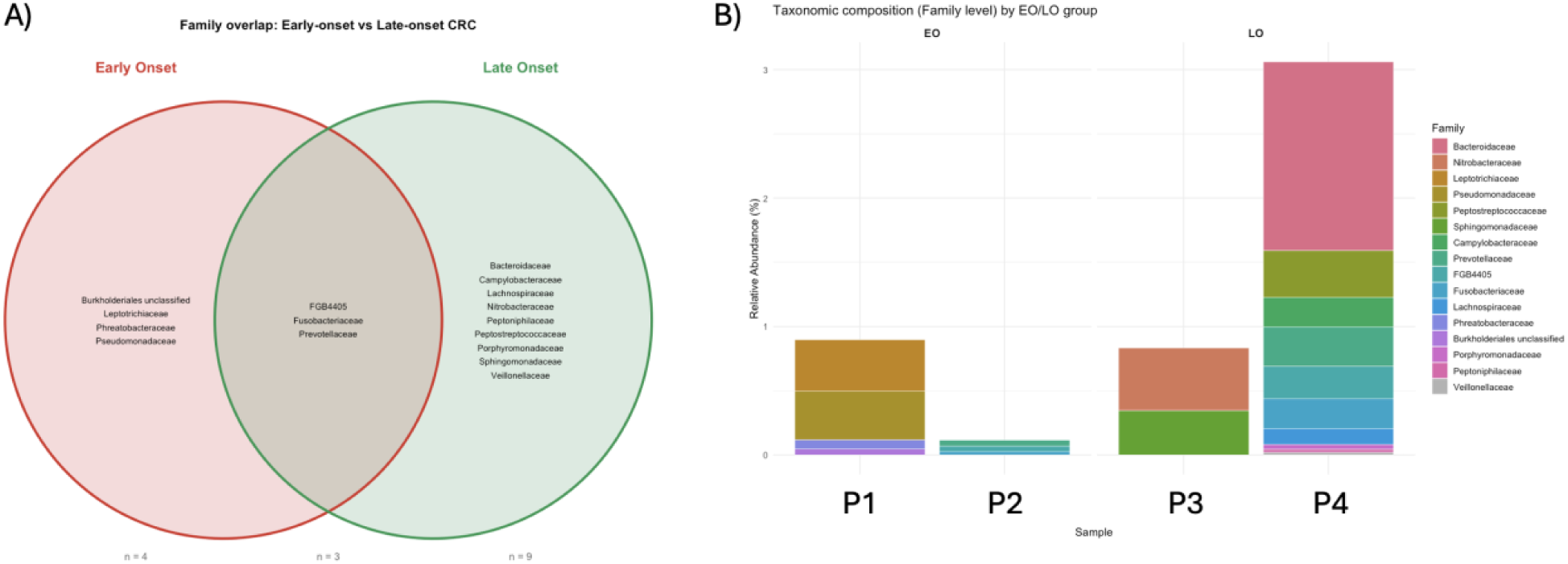
Family-Level Characterization of Tumor-Colonizing Microbiota in Early- and Late-Onset Colorectal Cancer Among Hispanic/Latino (H/L) Patients. (A) Venn diagram illustrating the overlap and unique distribution of bacterial families identified in tumor-colonizing microbiota. (B) Stacked bar plots showing the relative abundance of bacterial families in H/L colorectal tumors. Each bar represents an individual tumor, and colors denote bacterial families identified.

A total of 16 bacterial families were identified across the four tumors (Fig. 1A). Family-level composition differed markedly between EOCRC and LOCRC. EOCRC tumors contained four exclusive bacterial families (Eubacteriaceae, Paenibacillaceae, Pseudomonadaceae, and one unclassified family) whereas nine families were unique to LOCRC, including Enterobacteriaceae, Corynebacteriaceae, Moraxellaceae, Nocardiaceae, Peptostreptococcaceae, Sphingomonadaceae, Sphingobacteriaceae, Veillonellaceae, and Weeksellaceae. In contrast, Lactobacillaceae, Fusobacteriaceae, and Prevotellaceae were detected in both age groups, representing a shared core tumor microbiota.

LOCRC harbored more than twice the number of unique bacterial families observed in EOCRC, indicating substantially greater microbial richness in tumors from older patients (Fig. 1A). Conversely, EOCRC exhibited a comparatively restricted family-level microbial composition, suggesting distinct tumor-associated microbial communities according to age at disease onset.

To further characterize these differences, we examined the relative abundance of bacterial families within individual tumors (Fig. 1B). Considerable inter-patient heterogeneity was observed; however, consistent age-associated patterns emerged. The EOCRC tumors displayed relatively simple microbial communities dominated by a limited number of bacterial families. One EOCRC tumor was enriched in Prevotellaceae and Leptotrichiaceae, whereas the second exhibited only low-abundance bacterial families, indicating relatively low taxonomic complexity.

In contrast, LOCRC tumors demonstrated substantially more complex microbial profiles. One LOCRC sample was enriched for Fusobacteriaceae together with Porphyromonadaceae, whereas the second contained a diverse community comprising Peptostreptococcaceae, Lachnospiraceae, Prevotellaceae, Fusobacteriaceae, Bacteroidaceae, Veillonellaceae, and several additional low-abundance families. The simultaneous detection of multiple anaerobic bacterial families highlights the polymicrobial nature of the colorectal TME.

Notably, Fusobacteriaceae, a bacterial family strongly associated with colorectal carcinogenesis, was identified in tumors from both EOCRC and LOCRC, whereas several anaerobic families previously implicated in CRC were predominantly detected in LOCRC. Together, the family-level overlap (Fig. 1A) and abundance profiles (Fig. 1B) consistently demonstrate that LOCRC harbor more taxonomically diverse tumor-colonizing microbiota than early-onset tumors. Although this pilot study includes only four representative H/L tumors, these findings provide preliminary evidence that age at diagnosis is associated with differences in the structural composition of the tumor microbiome and establish a framework for larger studies integrating microbial profiling with host genomics, immune characteristics, and clinical outcomes.

#### 2.2.2 Genus-Level Taxonomic Composition

To further characterize age-associated differences in the tumor microbiome, we performed genus-level taxonomic profiling on WES data from H/L colorectal tumors (Fig. 2A,B). Genus-level analysis identified 18 bacterial genera, revealing both conserved and age-specific microbial communities within the colorectal TME.

**Figure 2.**
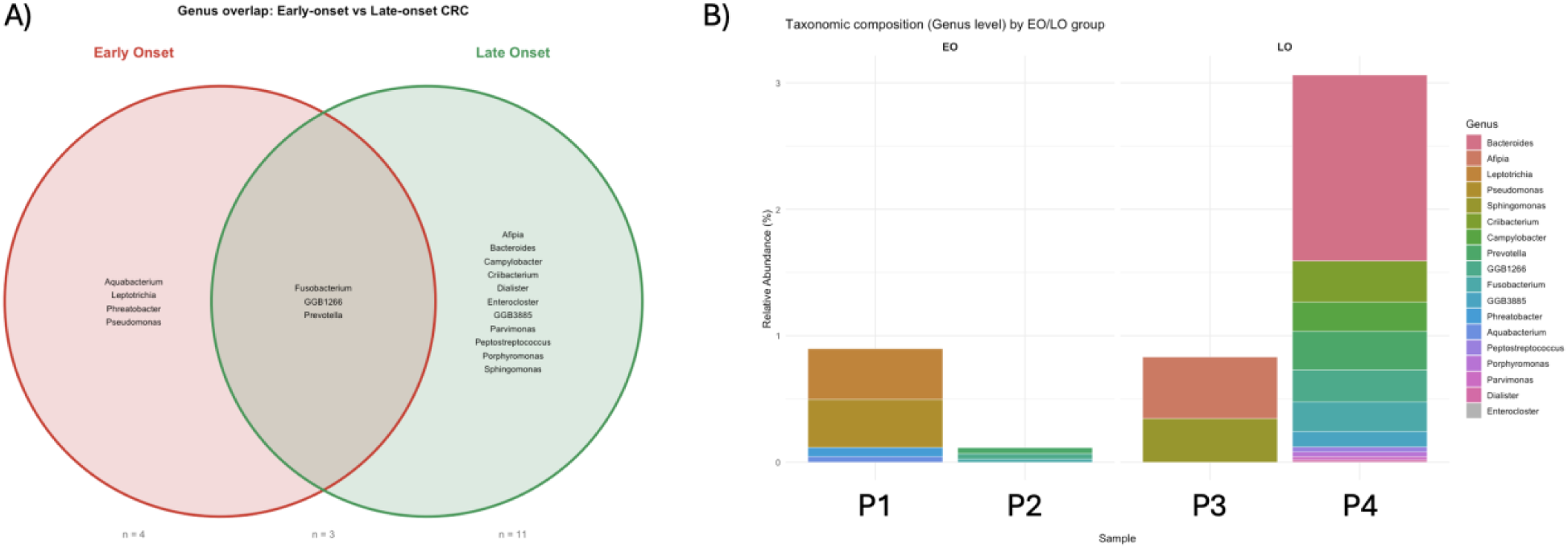
Genus-Level Characterization of Tumor-Colonizing Microbiota in Early- and Late-Onset Colorectal Cancer Among Hispanic/Latino Patients. (A) Venn diagram illustrating the overlap and unique distribution of bacterial genera identified in four representative colorectal tumors. (B) Stacked bar plots showing the relative abundance of bacterial genera identified. Each bar represents an individual tumor sample.

Comparison of EOCRC and LOCRC demonstrated marked differences in microbial composition (Fig. 2A). EOCRC tumors contained four exclusive genera (Aquabacterium, Leptotrichia, Paenibacillus, and Pseudomonas) whereas LOCRC tumors harbored 11 unique genera, including Herbaspirillum, Enterobacter, Cloacibacterium, Delftia, Elizabethkingia, Pedobacter, Peptoniphilus, Porphyromonas, and Sphingomonas. In contrast, Cutibacterium, Fusobacterium, and Prevotella were detected in both age groups, representing a conserved core tumor microbiota.

Overall, LOCRC exhibited nearly three-fold more unique bacterial genera than EOCRC, indicating substantially greater genus-level microbial diversity in tumors from older patients (Fig. 2A). These findings extend the family-level analyses (Fig. 1) and demonstrate that age-associated differences in microbial composition are preserved across multiple taxonomic levels.

To further examine microbial composition within individual tumors, we compared the relative abundance of bacterial genera in each representative sample (Fig. 2B). Although considerable inter-patient heterogeneity was observed, consistent age-associated patterns emerged. EOCRC tumors displayed relatively simple microbial communities dominated by fewer bacterial genera. One EOCRC tumor was enriched in Leptotrichia and Pseudomonas, with lower relative abundances of Prevotella and Fusobacterium, whereas the second EOCRC tumor contained only a limited number of low-abundance genera, indicating relatively low microbial complexity.

In contrast, LOCRC tumors demonstrated substantially more complex microbial communities. One LOCRC sample was enriched in Fusobacterium together with Porphyromonas, whereas the second exhibited the highest genus-level diversity among all tumors and was dominated by Bacteroides, with additional contributions from Prevotella, Cutibacterium, Sphingomonas, Acinetobacter, Delftia, Parvimonas, and Elizabethkingia. The coexistence of multiple anaerobic bacterial genera highlights the polymicrobial nature of the colorectal TME.

Among the detected microorganisms, Fusobacterium represented one of the most biologically relevant genera because of its well-established role in colorectal carcinogenesis. Previous studies have shown that Fusobacterium nucleatum promotes epithelial invasion, activation of WNT/β-catenin signaling, immune evasion, and metastatic progression. Consistent with these observations, Fusobacterium was detected in tumors from both age groups but was more abundant in LOCRC, where it co-occurred with additional anaerobic genera including Porphyromonas, Parvimonas, and Prevotella, suggesting the presence of cooperative microbial communities associated with tumor progression.

Similarly, the predominance of Bacteroides in one LOCRC tumor is noteworthy because this genus includes enterotoxigenic Bacteroides fragilis, a recognized CRC-associated pathogen capable of inducing epithelial barrier dysfunction, chronic inflammation, DNA damage, and activation of oncogenic signaling pathways. Although species-level characterization is presented in the following section, enrichment of the Bacteroides genus further supports its role as a major component of tumor-associated microbial ecosystems.

The genus-level overlap (Fig. 2A) and abundance profiles (Fig. 2B) consistently demonstrate that LOCRC harbor greater genus-level microbial richness and complexity than early-onset tumors. Despite the exploratory nature of this pilot study, the concordant patterns observed across family- and genus-level analyses provide preliminary evidence that EOCRC and LOCRC develop within distinct tumor-associated microbial ecosystems. These findings establish a foundation for future studies integrating microbial profiling with host genomics, immune landscapes, and clinical outcomes to identify microbiome-based biomarkers for precision oncology in H/L CRC.

#### 2.2.3 Species-Level Taxonomic Composition

To determine whether age-associated differences in the tumor microbiome extended to the highest taxonomic resolution, we performed species-level microbial profiling on WES data (Fig. 3A,B). Species-level analysis identified 20 bacterial species, revealing pronounced divergence in tumor-colonizing microbiota between EOCRC and LOCRC.

**Figure 3.**
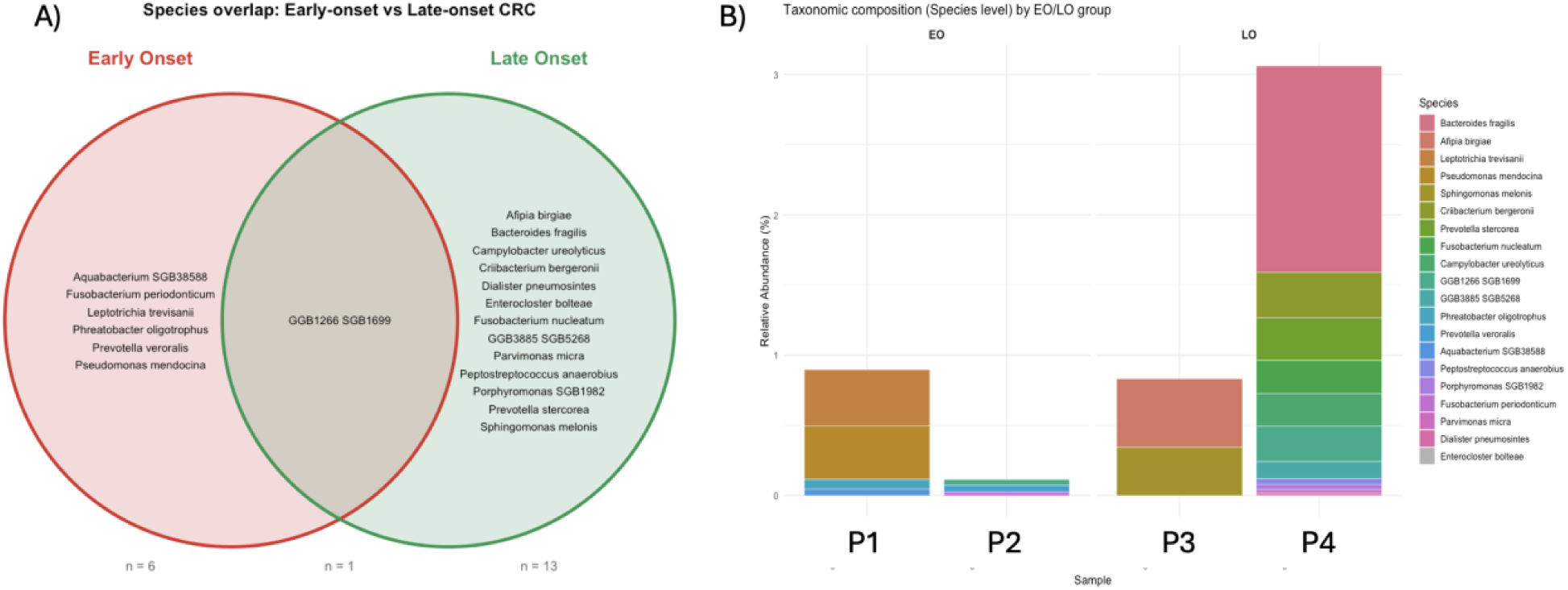
Species-Level Characterization of Tumor-Colonizing Microbiota in Early- and Late-Onset Colorectal Cancer Among Hispanic/Latino Patients. (A) Venn diagram illustrating the overlap and unique distribution of bacterial species identified in colorectal tumors. (B) Stacked bar plots showing the relative abundance of bacterial species identified. Each bar represents an individual tumor sample.

Comparison of both age groups demonstrated marked differences in species composition (Fig. 3A). EOCRC tumors contained six exclusive bacterial species, including Fusobacterium sp. BG528868, Leptotrichia hongkongensis, Paenibacillus oligotrophicus, Prevotella veroralis, Pseudomonas mendocina, and one additional species classified within the Tumebacillus/Flavobacterium group. In contrast, LOCRC tumors harbored 13 unique bacterial species, including Afipia birgiae, Enterobacter ludwigii, Elizabethkingia ursingii, Cloacibacterium normanense, Dialister pneumosintes, Elizabethkingia bruuniana, Fusobacterium nucleatum, Parvimonas micra, Porphyromonas asaccharolytica, Prevotella stercorea, and Sphingomonas melonis. Only a single unidentified bacterial species (GGB1266/GGB1669) was shared between EOCRC and LOCRC, indicating minimal overlap at the species level.

The marked reduction in shared taxa compared with the family- and genus-level analyses suggests that age-associated differences become increasingly pronounced with higher taxonomic resolution. Consistent with the preceding analyses, LOCRC exhibited substantially greater species-level diversity than EOCRC, supporting the presence of distinct tumor-associated microbial ecosystems according to age at disease onset.

Species-level abundance profiles further highlighted these differences (Fig. 3B). Although considerable inter-patient heterogeneity was observed, consistent age-associated patterns emerged. EOCRC tumors displayed relatively simple microbial communities dominated by a limited number of bacterial species. One EOCRC tumor was enriched in Leptotrichia trevisanii and Pseudomonas mendocina, with lower abundances of Prevotella veroralis, Fusobacterium sp. BG528868, and Aquabacterium spp., whereas the second EOCRC tumor contained only a small number of low-abundance bacterial species.

In contrast, LOCRC tumors exhibited substantially greater species richness and taxonomic complexity. One LOCRC tumor was enriched in Fusobacterium nucleatum together with Porphyromonas species, whereas the second displayed the highest species diversity among all tumors and was dominated by Bacteroides fragilis. Additional abundant species included Prevotella stercorea, Parvimonas micra, Dialister pneumosintes, Sphingomonas melonis, Afipia birgiae, Campylobacter ureolyticus, and Enterobacter species. The simultaneous detection of numerous anaerobic and facultative anaerobic bacteria underscores the polymicrobial nature of the colorectal TME.

Among the identified microorganisms, Fusobacterium nucleatum represented one of the most biologically relevant findings because of its well-established role in colorectal carcinogenesis. Previous studies have demonstrated that F. nucleatum promotes epithelial invasion, activation of WNT/β-catenin signaling, immune evasion, biofilm formation, and metastatic progression. Consistent with these observations, F. nucleatum was predominantly detected in LOCRC tumors, where it co-occurred with additional CRC-associated anaerobes including Parvimonas micra, Porphyromonas asaccharolytica, and Dialister pneumosintes, suggesting cooperative microbial networks associated with tumor progression.

Similarly, the predominance of Bacteroides fragilis in one LOCRC tumor is noteworthy because enterotoxigenic strains of this species have been implicated in epithelial barrier disruption, chronic inflammation, DNA damage, and activation of oncogenic signaling pathways. Although strain-level analyses were beyond the scope of this study, its enrichment further supports the contribution of Bacteroides to colorectal tumor-associated microbial ecosystems.

The species-level overlap (Fig. 3A) and abundance profiles (Fig. 3B) extend the family- and genus-level analyses by demonstrating that late-onset CRC harbor substantially greater species-level diversity than early-onset tumors. The progressive reduction in shared taxa from family to genus to species further indicates that microbial divergence becomes increasingly apparent at higher taxonomic resolution. Although based on four representative H/L tumors, these concordant findings provide preliminary evidence that EOCRC and LOCRC develop within distinct intratumoral microbial ecosystems and establish a framework for future studies integrating species-level microbiome profiling with host genomics, immune landscapes, and clinical outcomes.

### 2.3. Functional Characterization of Tumor-Colonizing Microbiota in EOCRC and LOCRC

To determine whether the taxonomic differences observed between EOCRC and LOCRC were accompanied by alterations in microbial metabolic potential, we performed functional pathway profiling on WES data from H/L colorectal tumors, including EOCRC and LOCRC samples (Fig. 4A,B). Functional profiling enabled inference of the metabolic capabilities encoded by the intratumoral microbial communities and assessment of whether age-associated taxonomic variation translated into functional divergence.

**Figure 4.**
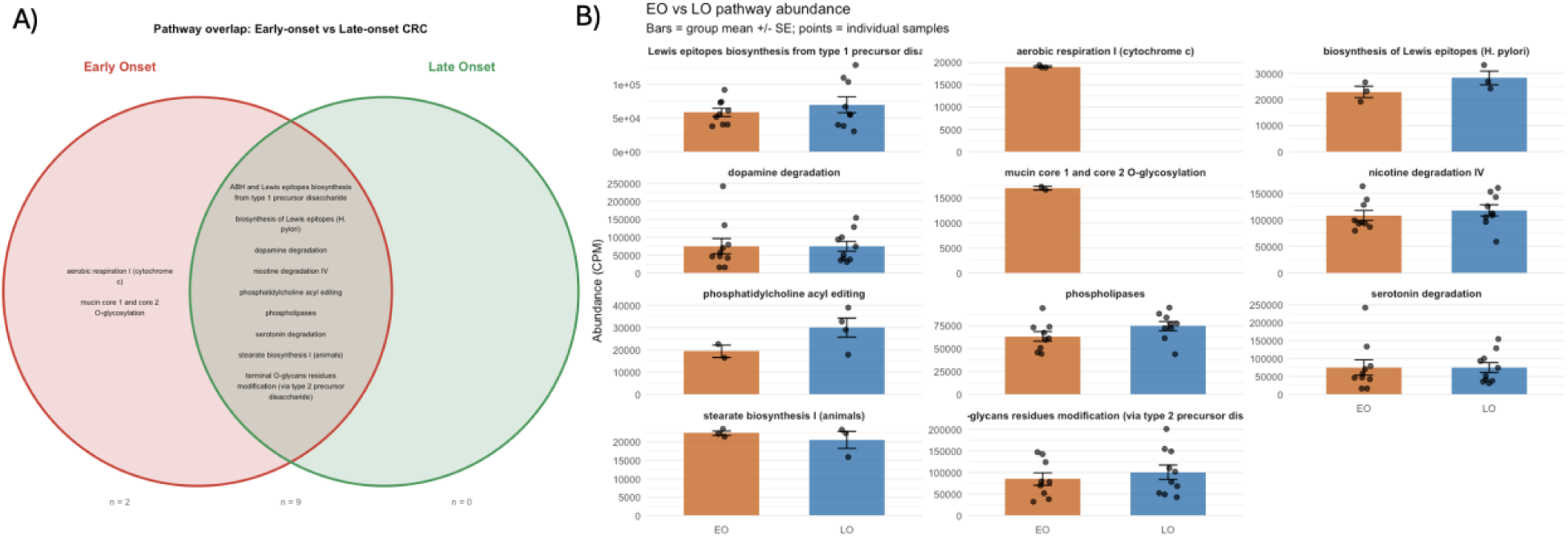
Functional Characterization of Tumor-Colonizing Microbiota in Early- and Late-Onset Colorectal Cancer Among Hispanic/Latino Patients. (A) Venn diagram illustrating the overlap of predicted microbial metabolic pathways inferred. (B) Comparative abundance of predicted microbial metabolic pathways in EOCRC and LOCRC tumors. Bar plots represent the mean pathway abundance for each age group, with individual data points corresponding to individual tumors.

Overall, 11 predicted microbial metabolic pathways were identified across the study cohort (Fig. 4A). In contrast to the marked differences observed at the family, genus, and species levels, microbial functional profiles were highly conserved between EOCRC and LOCRC. Specifically, nine pathways were shared between both age groups, two pathways were unique to EOCRC, and no pathways were exclusive to LOCRC. The EOCRC-specific pathways were associated with aromatic compound metabolism and mucin-associated glycan degradation, suggesting limited age-specific metabolic adaptations within tumors from younger patients.

The shared pathways encompassed essential microbial processes, including energy metabolism, amino acid biosynthesis, carbohydrate utilization, nucleotide metabolism, phospholipid metabolism, cofactor and vitamin biosynthesis, and central carbon metabolism, indicating that core microbial functions are maintained despite substantial differences in community composition. Notably, the absence of LOCRC-specific pathways suggests that the increased microbial diversity observed in late-onset tumors reflects expansion of taxonomic diversity rather than acquisition of novel metabolic capabilities.

To further evaluate pathway activity, we compared the relative abundance of predicted microbial functions between EOCRC and LOCRC (Fig. 4B). The abundance of most shared pathways was remarkably similar across both groups, further supporting conservation of microbial metabolic function despite pronounced taxonomic divergence. Pathways involved in aerobic respiration (cytochrome c), L-methionine salvage, phosphatidylcholine biosynthesis, phospholipid metabolism, acetate degradation, tetrapyrrole (heme) biosynthesis, and Lewis antigen-associated glycan biosynthesis exhibited comparable abundance in EOCRC and LOCRC, indicating preservation of fundamental metabolic activities within the TME.

Although average pathway abundances remained similar, several pathways demonstrated greater inter-patient variability, particularly among LOCRC tumors. Pathways involved in adenosine degradation and serotonin degradation showed increased heterogeneity, suggesting that host-microbiome metabolic interactions may vary substantially between individual tumors. Consistent with the greater taxonomic diversity observed in LOCRC, functional variability was also modestly increased across several metabolic pathways, despite preservation of the overall functional landscape.

These findings demonstrate a striking contrast between taxonomic diversity and functional conservation. While EOCRC and LOCRC harbor markedly different tumor-colonizing microbial communities, their predicted metabolic capabilities remain largely preserved. This pattern is consistent with functional redundancy, whereby phylogenetically distinct microbial taxa encode overlapping metabolic functions and collectively maintain essential biological processes within the colorectal TME. Although this pilot study is limited to four representative H/L tumors, it provides proof-of-concept that the intratumoral microbiome exhibits high taxonomic plasticity but substantial functional conservation, establishing a foundation for future multi-omics studies integrating microbial function with host genomics, immune landscapes, metabolomics, and clinical outcomes.

## 3. Discussion

The incidence of EOCRC continues to increase worldwide, particularly among H/L populations, yet the biological mechanisms underlying this emerging disease remain incompletely understood. Although growing evidence implicates the gut microbiome as an important contributor to colorectal carcinogenesis, relatively little is known about the composition of tumor-colonizing microbial communities in EOCRC, particularly within ancestrally diverse populations. In this pilot study, we performed an integrative characterization of the tumor microbiome in H/L patients with EOCRC and LOCRC using microbial profiling derived from WES sequencing data. Across multiple taxonomic levels, our findings demonstrate that tumor-associated microbial communities differ according to age at diagnosis, with LOCRC exhibiting substantially greater microbial richness and taxonomic diversity than EOCRC. Despite these marked compositional differences, predicted microbial metabolic functions remained remarkably conserved, suggesting that distinct microbial communities converge on similar biological programs within the colorectal TME.

One of the most consistent observations across the present study was the progressive increase in microbial divergence from the family to genus and species levels. While several bacterial families and genera were shared between EOCRC and LOCRC, overlap decreased substantially at higher taxonomic resolution, where only a single microbial species was detected in both age groups. This progressive reduction in shared taxa suggests that age-associated microbial differences become increasingly evident as taxonomic resolution improves. Such findings are consistent with recent metagenomic studies demonstrating that species-level analyses often reveal biologically relevant microbial signatures that remain obscured at broader taxonomic classifications. Our data therefore support the concept that high-resolution microbial profiling may be necessary to fully understand microbiome-associated heterogeneity in CRC.

Another important finding was the consistently greater microbial diversity observed in LOCRC. At every taxonomic level examined, LOCRC tumors contained more unique bacterial families, genera, and species than EOCRC. Increased microbial richness within colorectal tumors has previously been associated with chronic inflammation, prolonged environmental exposure, dietary influences, and cumulative alterations of the intestinal microenvironment that accompany aging. The broader microbial ecosystems observed in LOCRC may therefore reflect the longer temporal evolution of host-microbe interactions before malignant transformation. Conversely, the relatively restricted microbial communities identified in EOCRC raise the possibility that younger patients develop CRC through distinct biological mechanisms requiring fewer microbial taxa or more specialized microbial networks.

Despite these differences, our analyses also identified a conserved core tumor microbiota shared between EOCRC and LOCRC. Members of the Fusobacteriaceae, Prevotellaceae, and Lactobacillaceae families, together with the genera Fusobacterium, Prevotella, and Cutibacterium, were detected across both age groups, suggesting that certain microorganisms participate broadly in colorectal tumor biology regardless of patient age. Among these taxa, Fusobacterium remains the most compelling candidate because of its well-established role in colorectal carcinogenesis. Numerous experimental studies have demonstrated that Fusobacterium nucleatum promotes epithelial invasion, activation of WNT/β-catenin signaling, suppression of anti-tumor immune responses, induction of inflammatory cytokines, chemoresistance, and metastatic dissemination. The observation that Fusobacterium persisted across multiple taxonomic levels in our cohort further supports its role as a central component of the colorectal tumor microbiome.

In contrast, several microorganisms appeared to distinguish LOCRC from EOCRC. LOCRC tumors demonstrated enrichment of Bacteroides, Porphyromonas, Parvimonas, Dialister, Enterobacter, and Sphingomonas, together with species including Bacteroides fragilis, Fusobacterium nucleatum, Parvimonas micra, Porphyromonas asaccharolytica, and Prevotella stercorea. Many of these organisms have previously been implicated in chronic inflammation, epithelial barrier dysfunction, immune modulation, biofilm formation, and metabolic reprogramming within colorectal tumors. In particular, enterotoxigenic Bacteroides fragilis produces the B. fragilis toxin capable of disrupting epithelial integrity, inducing DNA damage, and activating oncogenic signaling pathways. Likewise, Parvimonas micra and Porphyromonas species have emerged as reproducible components of CRC-associated polymicrobial biofilms and have been associated with disease progression and poor clinical outcomes. The enrichment of these microorganisms in LOCRC suggests that microbial cooperation within complex anaerobic communities may become increasingly important during colorectal tumor evolution in older patients.

Conversely, EOCRC tumors contained several unique bacterial taxa, including Leptotrichia, Pseudomonas, Paenibacillus, and Aquabacterium, together with species such as Leptotrichia hongkongensis, Pseudomonas mendocina, Paenibacillus oligotrophicus, and Prevotella veroralis. Although these organisms have not been consistently associated with CRC, several have been linked to mucosal microbial communities or opportunistic colonization. Their presence exclusively in EOCRC raises the possibility that younger patients develop tumors within microbial ecosystems that differ fundamentally from those observed in older individuals. Whether these microorganisms contribute directly to carcinogenesis or instead reflect age-associated ecological differences remains to be determined.

Perhaps the most intriguing observation of the present study is the apparent disconnect between microbial composition and predicted microbial function. Although taxonomic analyses consistently demonstrated greater microbial diversity in LOCRC, functional profiling revealed that the overwhelming majority of microbial metabolic pathways were shared between EOCRC and LOCRC. Core pathways involved in oxidative respiration, phospholipid metabolism, amino acid biosynthesis, carbon metabolism, and cofactor biosynthesis were highly conserved, whereas only two predicted pathways were uniquely detected in EOCRC and none were exclusive to LOCRC. These findings strongly support the concept of functional redundancy, whereby phylogenetically distinct microorganisms encode overlapping metabolic capabilities that collectively preserve essential biological functions within the TME. Functional redundancy represents a common ecological principle within complex microbial ecosystems and may explain why distinct microbial communities produce similar physiological effects despite substantial taxonomic variation.

From a translational perspective, our findings suggest that microbial composition and microbial function provide complementary biological information. Taxonomic profiling may identify biomarkers useful for distinguishing EOCRC from LOCRC or for understanding disease heterogeneity, whereas functional analyses may reveal conserved metabolic pathways that represent more stable therapeutic targets. Integrating both taxonomic and functional information with host genomic, transcriptomic, immune, and metabolomic data may therefore provide a more comprehensive understanding of host-microbiome interactions and improve patient stratification for precision oncology.

An important strength of this study is the exclusive focus on H/L patients, an understudied population that experiences a disproportionate increase in EOCRC incidence. Most microbiome studies have been conducted in predominantly European or Asian populations, limiting generalizability across ancestries. Because host genetics, environmental exposures, dietary habits, and socioeconomic determinants of health influence microbial composition, characterization of tumor-associated microbiota within H/L populations represents an important step toward equitable precision medicine. Although exploratory, our findings demonstrate the feasibility of deriving meaningful microbial information from clinically generated WES datasets and provide an initial framework for future ancestry-specific microbiome investigations.

Several limitations should be acknowledged. First, this is a pilot study involving only four representative tumors, precluding formal statistical comparisons and limiting the ability to evaluate associations with clinicopathologic variables, treatment response, or survival. Second, microbial profiling was performed using WES sequencing, which was originally designed for host genomic analysis rather than microbiome characterization. Consequently, species detection is limited by sequencing depth and capture bias compared with dedicated shotgun metagenomic sequencing. Third, microbial function was inferred computationally rather than measured directly using metatranscriptomics or metabolomics. Fourth, the cross-sectional design precludes inference regarding temporal relationships or causal contributions of individual microorganisms to tumor initiation or progression. Finally, the absence of matched normal tissue, stool microbiome, immune profiling, and longitudinal clinical follow-up limits biological interpretation.

Despite these limitations, the concordant findings observed across family-, genus-, species-, and functional-level analyses provide compelling preliminary evidence that EOCRC and LOCRC develop within distinct tumor-associated microbial ecosystems while preserving a largely conserved functional landscape. These observations support a model in which age-associated colorectal carcinogenesis is accompanied by remodeling of microbial community structure without major disruption of essential microbial metabolic programs.

An important future direction arising from this work is the development of dedicated artificial intelligence (AI) agents [30–32] for CRC microbiome research. The rapidly expanding volume of microbiome, genomic, transcriptomic, metabolomic, immune, and clinical data presents significant challenges for comprehensive data integration and biological interpretation. AI-driven microbiome agents, such as AI-HOPE-Microbiome developed by the Velazquez-Villarreal lab at City of Hope [33], could systematically integrate tumor-associated microbial profiles with host molecular characteristics, clinicopathologic features, and continuously evolving scientific literature to accelerate hypothesis generation, biomarker discovery, and mechanistic interpretation. Such platforms could identify age-specific microbial signatures, prioritize candidate microbial biomarkers, and uncover functional interactions between the microbiome and the TME that may not be readily apparent through conventional analytical approaches. As larger multi-center and multi-omics datasets become available, a CRC microbiome AI agent could serve as a scalable decision-support and research platform, facilitating reproducible analyses and accelerating the translation of microbiome discoveries into precision oncology, particularly for understudied populations such as H/L patients.

In conclusion, this study provides one of the first characterizations of tumor-colonizing microbiota in H/L patients with EOCRC and LOCRC. Our findings demonstrate greater microbial richness and taxonomic complexity in LOCRC, identify conserved microbial taxa shared across age groups, and reveal extensive functional redundancy despite pronounced compositional differences. Together, these results highlight the tumor microbiome as an additional layer of biological heterogeneity in CRC and establish a proof-of-concept for integrating tumor microbiome profiling into precision oncology. Future studies incorporating larger multicenter H/L cohorts, shotgun metagenomics, metatranscriptomics, metabolomics, spatial transcriptomics, host immune profiling and using autonomous AI-agents will be essential to validate these findings, identify microbiome-based biomarkers, and determine whether modulation of the tumor microbiome can improve prevention, early detection, and treatment strategies for CRC.

## 4. Methods

### Patient Cohort

This retrospective pilot study included four H/L patients diagnosed with colorectal adenocarcinoma from City of Hope Comprehensive Cancer Center (COH). WES data, and corresponding clinicopathological information were obtained through institutional databases and precision oncology programs under Institutional Review Board (IRB)-approved protocols.

Patients were stratified according to age at diagnosis into EOCRC and LOCRC. Clinical variables included age at diagnosis, sex, tumor stage, primary tumor location, and pathological characteristics. For this proof-of-concept analysis, four primary colorectal tumors were selected, including two EOCRC and two LOCRC cases, representing both sexes and disease stages.

### Tumor Samples and Whole-Exome Sequencing

Raw sequencing data underwent institutional quality-control procedures, including assessment of sequencing quality, read depth, mapping efficiency, and duplicate read removal. Reads were aligned to the GRCh38 human reference genome using standard bioinformatics pipelines for somatic genomic analyses. The aligned BAM files generated from these analyses served as the source for subsequent microbial profiling.

### Identification of Tumor-Colonizing Microbiota

To characterize tumor-associated microbial communities, non-human sequencing reads were inferred from WES data following removal of host-derived sequences. Microbial taxonomic profiling was performed using MetaPhlAn, which identifies microorganisms based on clade-specific marker genes and enables direct taxonomic assignment from shotgun sequencing data without the need for targeted microbial sequencing.

Microbial communities were characterized at the family, genus, and species taxonomic levels. Taxa identified in EOCRC and LOCRC tumors were compared to evaluate differences in microbial composition associated with age at disease onset. Overlap between age groups was visualized using Venn diagrams, while taxonomic abundance within individual tumors was represented using stacked bar plots.

Because WES is optimized for host genomic characterization rather than microbiome analysis, the present study was designed as an exploratory analysis to identify robust tumor-colonizing microbial signatures detectable from tumor-derived sequencing data.

### Functional Profiling of Tumor-Colonizing Microbiota

To evaluate whether differences in microbial composition were accompanied by alterations in microbial metabolic potential, functional pathway profiling was performed using the MetaPhlAn/HUMAnN analytical framework. HUMAnN reconstructs microbial metabolic pathways from taxonomic profiles and estimates the relative abundance of predicted microbial functions.

Predicted pathways were compared between EOCRC and LOCRC to assess conservation or divergence of microbial metabolic activities. Functional overlap between age groups was summarized using Venn diagrams, and pathway abundances were compared using descriptive analyses of individual tumors.

### Clinical Data Collection

Clinical and pathological information was extracted from institutional research databases. Variables included age at diagnosis, sex, American Joint Committee on Cancer (AJCC) pathological stage, and tumor location. Patients were categorized as EOCRC or LOCRC based on age at diagnosis, and descriptive comparisons were performed between groups.

### Data Visualization

Taxonomic overlap at the family, genus, species, and functional pathway levels was visualized using Venn diagrams. Relative microbial abundances within individual tumors were illustrated using stacked bar plots generated in R. Functional pathway abundances were summarized using bar graphs displaying individual tumors and group-level trends.

### Statistical Analysis

Given the exploratory nature of this pilot study and the limited sample size (n = 4), analyses were primarily descriptive. Taxonomic composition and predicted microbial functions were compared qualitatively between EOCRC and LOCRC to identify conserved and age-specific microbial signatures. Continuous variables are presented descriptively, and categorical variables are summarized as frequencies and percentages. Statistical hypothesis testing was not performed because the study was designed as a proof-of-concept investigation rather than a powered comparative analysis.

## Data Availability

All data produced in the present study are available upon reasonable request to the authors.

